# Severe COVID-19 and Diabetes - A Retrospective Cohort Study from Three London Teaching Hospitals

**DOI:** 10.1101/2020.08.07.20160275

**Authors:** Chioma Izzi-Engbeaya, Walter Distaso, Anjali Amin, Wei Yang, Oluwagbemiga Idowu, Julia S Kenkre, Ronak J Shah, Evelina Woin, Christine Shi, Nael Alavi, Hala Bedri, Niamh Brady, Sophie Blackburn, Martina Leczycka, Sanya Patel, Elizaveta Sokol, Edward Toke-Bjolgerud, Ambreen Qayum, Mariana Abdel-Malek, David C D Hope, Nick S Oliver, Vasiliki Bravis, Shivani Misra, Tricia M Tan, Neil Hill, Victoria Salem

## Abstract

Patients with diabetes mellitus admitted to hospital with COVID-19 caused by infection with the novel coronavirus (SARS-CoV-2) have poorer outcomes. However, the drivers for this are not fully elucidated. We performed a retrospective cohort study, including detailed pre-hospital and presenting clinical and biochemical factors of 889 patients diagnosed with COVID-19 in three constituent hospitals of a large London NHS Trust. 62% of patients with severe COVID-19 were of non-White ethnic backgrounds and the prevalence of diabetes was 38%. 323 (36%) patients met the primary outcome of death or admission to the intensive care unit (ICU) within 30 days of diagnosis. Male gender, advancing age and the Clinical Frailty Scale, an established measure of multimorbidity, independently predicted poor outcomes on multivariate analysis. Diabetes did not confer an independent risk for adverse outcomes in COVID-19, although patients with diabetes and ischaemic heart disease were at particular risk. Additional risk factors which significantly and independently associated with poorer outcomes in patients with diabetes were age, male gender and lower platelet count. Antiplatelet medication was associated with a lower risk of death/ICU admission and should be evaluated in randomised clinical trials amongst high risk patient groups.

## Introduction

The World Health Organisation (WHO) announced a global pandemic of COVID-19 Severe Acute Respiratory Syndrome (SARS) caused by the novel coronavirus (SARS-CoV-2) on 11^th^ March 2020. Wide-ranging restrictions aimed at reducing spread of the virus (“lockdown measures”) were introduced in the United Kingdom (UK) on 23^rd^ March 2020. Around this time, London hospitals experienced the first surge of cases. By late June in the UK (28^th^ June 2020), there were 311,151 confirmed cases and 43,550 deaths. On 7^th^ July 2020, the UK Office for National Statistics reported that excess deaths had fallen below the 5-year average for the second consecutive week since the coronavirus pandemic was announced^1^, marking the end of the first wave of infections in England. However, there remain local areas of increasing infection rates across the country and a risk of future upsurges^2^. Therefore, understanding clinical correlates with disease severity, from the point of view of hospital triaging and advice on shielding, remains of paramount importance.

In February 2020, the first cohort studies from Wuhan were reporting clinical outcomes of patients treated for COVID-19^3-7^. These earliest studies reported age and male gender as major risk factors for poorer outcomes. Patients with comorbidities, most commonly hypertension and ischaemic heart disease, were over-represented in hospital or intensive care unit (ICU) admissions^3,5,6^, although it was not immediately clear to what extent these were independent of advancing age.

Early on it was noted that the prevalence of diabetes in patients admitted to ICU for severe COVID-19 (typically 20-30%^5,7^) was much greater than the adult population prevalence, which is 6.8% in the UK^8^. Patients infected with the prior two established human coronavirus infections (SARS-CoV and Middle East Respiratory Syndrome, MERS-CoV) were more likely to experience more severe disease and die if they had a diagnosis of diabetes^9,10^. However, granularity on the way diabetes interacts with the natural history of COVID-19 has been slower to come and remains an area of intense concern for patients with diabetes^11^ and their healthcare providers.

Two major hospital cohort studies have provided some insight about the interaction between diabetes and COVID-19. Zhu et al concluded that good glycaemic control in the acute hospital setting in China was an important factor for better outcomes in patients with pre-existing type 2 diabetes, although it was difficult to ascertain whether this was confounded by the possibility that poorer glycaemia in hospital was itself a marker of a more severe inflammatory response or the decision to use corticosteroids^12^. In Europe, a French consortium published a multicentre observational study (CORONADO) looking at clinical features associated with disease severity in people with diabetes hospitalised for COVID-19. The CORONADO study reported that on multivariate analysis of patients with diabetes hospitalised for COVID-19, BMI, but not glycated haemoglobin (HbA1C), was positively and independently associated with poorer outcomes^13^.

Here we report on all patients treated as an inpatient for swab-positive COVID-19 in three West London teaching hospitals, under the umbrella of Imperial College Healthcare NHS Trust (ICHNT), during our peak period of infections. We focus on clinical and biochemical features associated with COVID-19 disease severity within the whole cohort as well as the subset of patients with a diagnosis of diabetes. Our data suggest that it is age and its implicit association with multimorbidity that predicts poorer outcome, with the Clinical Frailty Scale (CFS) being a good measure of this. Patients with diabetes are at risk of more severe coronavirus infection because of increased association with other medical conditions, particularly ischaemic heart disease.

## Results

### STUDY COHORT

We report on 889 consecutive adult patients with a confirmed (SARS-CoV-2 swab positive) diagnosis of COVID-19 admitted to three Central London hospitals within the same NHS Trust (ICHNT) between 6^th^ March 2020 and 22^nd^ April 2020. A total of 323 (36%) of these patients met the primary outcome of death or admission to the intensive care unit (ICU) within 30 days of diagnosis. A total of 110 patients were admitted to ICU, of which 101 required endotracheal intubation and 50 of these ICU-admitted patients died. In the cohort of patients studied here, 337/889 (38%) had a diagnosis of diabetes, 96% (324/337) of people with diabetes had Type 2 diabetes (including 4 patients who were diagnosed during their admission for COVID-19) and the remainder (13 patients) had Type 1 diabetes. The Study Flow Diagram is shown in Figure 1.

**Figure 1:**
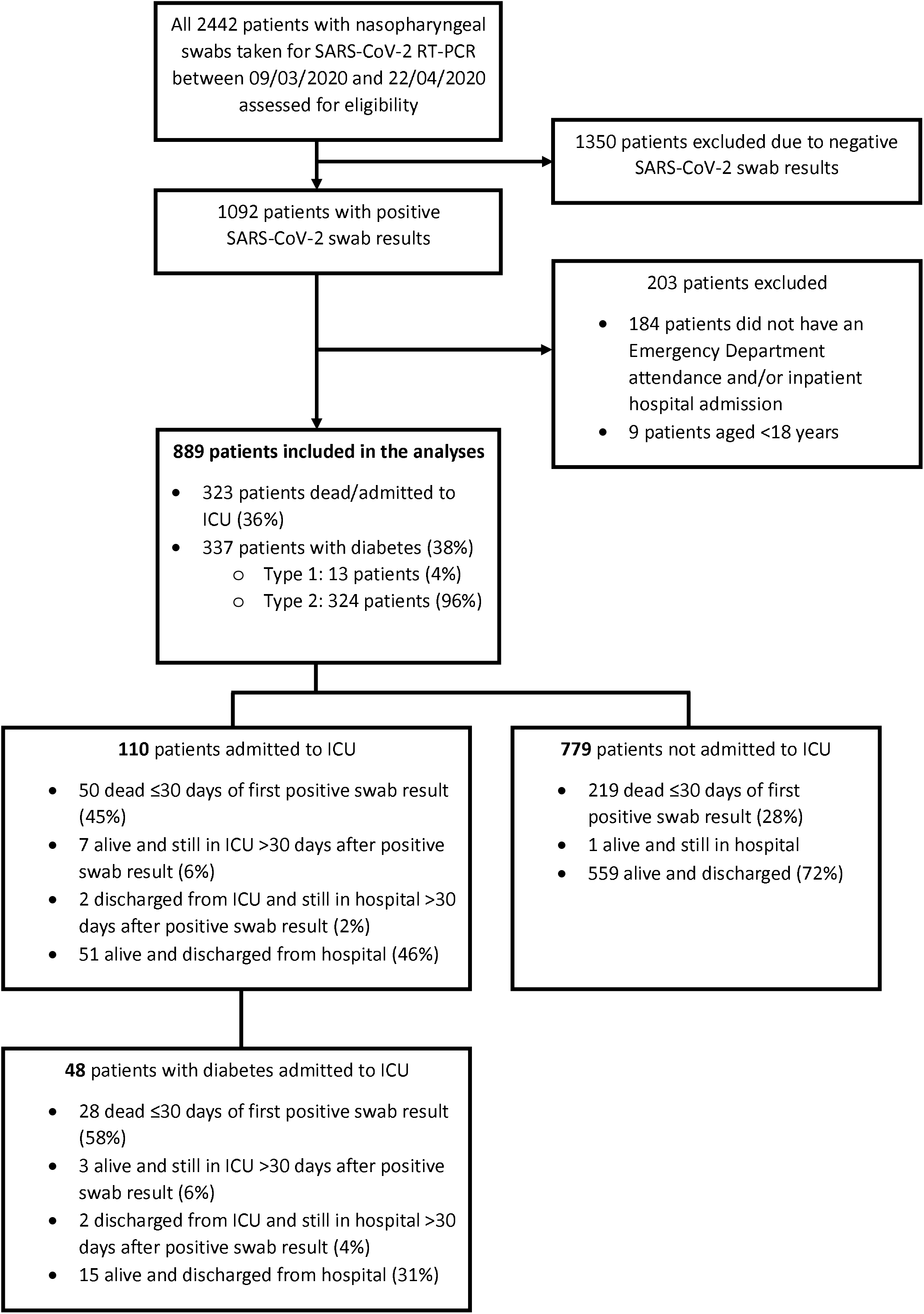
All 2442 patients who had nasopharyngeal swabs taken for reverse transcriptase polymerase chain reaction (RT-PCR) to detect the presence of SARS-CoV-2 (and diagnose COVID-19) between 9^th^ March 2020 and 22^nd^ April 2020 were assessed for eligibility. 1350 patients with negative swab results were excluded. Of 1092 patients with positive swab results, 203 were excluded (as they did not have an Emergency Department attendance and/or inpatient hospital admission, or they were aged less than 18 years); and 889 patients were included in the statistical analyses. eGFR = estimated glomerular filtration rate.

### FACTORS PRIOR TO HOSPITAL ADMISSION

The characteristics of the entire cohort are shown in Table 1. Of all patients treated for COVID-19 in our NHS Trust, the average (±SD) age was 65.8 (±17.5) years and 60% were men. The average (±SD) weight for men was 80.8 (±19.8) kg and for women 72.6 (±24.2) kg.

**Table 1.** – Demographic and Clinical Features of 889 Patients Treated for COVID-19 Patients in ICHNT Between March and April 2020 (Univariate Analysis for primary outcome of death or Intensive Care Unit admission within 30 days of swab positive diagnosis). 889 patients with confirmed (swab positive) COVID-19 were analysed and univariate odds ratios (OR) against a defined comparator are provided for our predefined primary outcome of death or ICU admission within 30 days of diagnosis. ACE inhibitor = angiotensin converting enzyme inhibitor, CBG = capillary blood glucose, CFS = Clinical Frailty Scale, COPD = Chronic obstructive pulmonary disease, DPPIV inhibitor = dipeptidyl peptidase-IV inhibitor, eGFR = estimated glomerular filtration rate, GLP-1RA = glucagons-like peptide-1 receptor agonist, HbA1c = glycated haemoglobin, SGLT2 inhibitor = sodium-glucose co-transporter-2 inhibitor. Significant p-values for odds ratios using Fishers’ exact test are shown in **bold**.

| WHOLE COHORT |  |  |  |  |  |  |
| --- | --- | --- | --- | --- | --- | --- |
| Clinical feature |  | Number of patients with data | Prevalence (n) | Odds ratio of primary outcome (OR) | 95% confidence interval (CI) | p-value |
| Female |  | 889 | 40% (355) | 0.69 | 0.52 to 0.91 | **0.01** |
| Age (years) |  | 889 |  |  |  |  |
|  | 18-49 |  | 18% (164) | 0.40 | 0.24 to 0.67 | **<0.0001** |
|  | 50-59 |  | 18% (157) | 1 | REFERENCE |  |
|  | 60-69 |  | 17% (147) | 1.11 | 0.74 to 1.84 | 0.55 |
|  | 70-79 |  | 22% (193) | 1.33 | 0.86 to 2.03 | 0.23 |
|  | >80 |  | 26% (228) | 1.41 | 0.92 to 2.15 | 0.21 |
| Weight (kg) |  | 693 |  |  |  |  |
|  | <80 |  | 61% (420) | 1 | REFERENCE |  |
|  | ≥80 |  | 39% (273) | 1.15 | 0.84 to 1.57 | 0.42 |
| Ethnicity |  | 865 |  |  |  |  |
|  | White |  | 38% (327) | 1 | REFERENCE |  |
|  | Black |  | 17% (146) | 1.21 | 0.81 to 1.81 | 0.40 |
|  | South Asian |  | 11% (97) | 1.04 | 0.64 to 1.66 | >0.9 |
|  | All non-White |  | 62% (538) | 1.35 | 1.01 to 1.81 | **0.04** |
| Smoking status |  | 484 |  |  |  |  |
|  | Current/ex-smoker |  | 189 | 1 | REFERENCE |  |
|  | Never smoked |  | 295 | 1.38 | 0.93 to 2.03 | 0.11 |
| Co-morbidities |  | 889 |  | *OR (co-morbidity vs without co-morbidity)* |  |  |
|  | Hypertension |  | 47% (418) | 2.41 | 1.82 to 3.20 | **<0.001** |
|  | Hyperlipidaemia |  | 33% (296) | 1.56 | 1.16 to 2.08 | **0.04** |
|  | Ischaemic heart disease |  | 16% (144) | 2.25 | 1.58 to 3.23 | **<0.001** |
|  | Stroke |  | 13% (120) | 0.98 | 0.66 to 1.46 | >0.9 |
|  | Heart failure |  | 10% (87) | 2.49 | 1.60 to 3.85 | **<0.001** |
|  | COPD |  | 9% (80) | 1.12 | 0.71 to 1.81 | 0.63 |
|  | Active cancer |  | 9% (79) | 1.15 | 0.72 to 1.86 | 0.62 |
| Pre-admission medications |  | 889 |  | *OR (taking specified medication vs not taking the medication)* |  |  |
|  | Statin |  | 42% (373) | 1.49 | 1.13 to 1.97 | **0.006** |
|  | Anti-platelet drug |  | 23% (204) |  | 0.92 to 1.75 | 0.14 |
|  | ACE inhibitor |  | 17% (151) | 1.57 | 1.10 to 2.23 | **0.013** |
|  | Angiotensin II receptor blocker |  | 13% (116) | 1.43 | 0.95 to 2.12 | 0.09 |
| CFS Score (1-9) |  | 756 |  |  |  |  |
|  | 1 - 2 |  | 40% (300) | 1 | REFERENCE |  |
|  | 3 - 4 |  | 32% (242) | 1.50 | 1.05 to 2.15 | **0.03** |
|  | 5 - 6 |  | 15% (110) | 1.75 | 1.20 to 2.52 | **0.003** |
|  | ≥7 |  | 13% (104) | 2.34 | 1.46 to 3.75 | **0.0004** |
| eGFR at time of COVID-19 diagnosis (mL/kg/min) |  | 880 |  |  |  |  |
|  | >90 |  | 27% (240) | 1 | REFERENCE |  |
|  | 60 - 90 |  | 31% (269) | 1.60 | 1.06 to 2.41 | **0.02** |
|  | 45 - 59 |  | 11% (95) | 2.35 | 1.40 to 3.91 | **0.001** |
|  | 30 - 44 |  | 10% (93) | 4.28 | 2.60 to 7.21 | **<0.0001** |
|  | 15 - 29 |  | 9% (78) | 3.02 | 1.78 to 5.16 | **<0.0001** |
|  | <15 |  | 12% (105) | 4.19 | 2.56 to 6.78 | **<0.0001** |
| Glucose at time of COVID-19 diagnosis |  | 652 |  |  |  |  |
| *WHOLE COHORT* | <10 mmol/L |  | 78% (508) | 1 | REFERENCE |  |
|  | ≥10 mmol/L |  | 22% (144) | 2.04 | 1.39 to 2.95 | **0.0002** |
|  |  | 394 |  |  |  |  |
| *PATIENTS WITHOUT DIABETES* | <10 mmol/L |  | 94% (369) | 1 | REFERENCE |  |
|  | ≥10 mmol/L |  | 6% (25) | 3.56 | 1.60 to 8.62 | **0.004** |
| **PATIENTS WITH DIABETES** |  |  |  |  |  |  |
| Diabetes diagnosis |  | 889 |  |  |  |  |
|  | Patients without diabetes |  | 62% (552) | 1 | REFERENCE |  |
|  | Patients with diabetes |  | 38% (337) | 1.68 | 1.27 to 2.12 | **0.0003** |
| Type of diabetes |  | 337 |  |  |  |  |
|  | Type 2 |  | 96% (324) | 1 | REFERENCE |  |
|  | Type 1 |  | 4% (13) | 0.37 | 0.11 to 1.27 | 0.16 |
| Duration of diabetes |  | 189 |  |  |  |  |
|  | <10 years |  | 47% (88) | 1 | REFERENCE |  |
|  | ≥10 years |  | 53% (101) | 1.82 | 1.03 to 3.34 | 0.054 |
| Diabetes medication at time of COVID-19 diagnosis |  | 337 |  | *OR (taking specified medication vs not taking the medication)* |  |  |
|  | Insulin |  | 31% (108) | 0.88 | 0.55 to 1.42 | 0.64 |
|  | GLP-1RA |  | 1% (5) | 0.52 | 0.09 to 2.60 | 0.66 |
|  | Metformin |  | 49% (169) | 1.14 | 0.74 to 1.76 | 0.58 |
|  | Sulphonylurea |  | 22% (74) | 0.85 | 0.50 to 1.45 | 0.59 |
|  | SGLT2 inhibitor |  | 7% (24) | 0.66 | 0.30 to 1.52 | 0.40 |
|  | DPPIV inhibitor |  | 27% (93) | 1.27 | 0.79 to 2.05 | 0.39 |
|  | Total number of diabetes medications |  |  |  |  |  |
|  | 0 |  | 18% (57) | 1 | REFERENCE |  |
|  | 1 |  | 41% (139) |  |  | 0.75 |
|  | 2 |  | 28% (96) |  |  | >0.9 |
|  | ≥3 |  | 13% (45) |  |  | >0.9 |
| HbA1c |  | 240 |  |  |  |  |
|  | <58 mmol/mol |  | 47% (112) | 1 | REFERENCE |  |
|  | ≥58 mmol/mol |  | 53% (128) | 1.44 | 0.85 to 2.39 | 0.19 |
| Glucose at time of COVID-19 diagnosis |  | 323 |  |  |  |  |
|  | <10 mmol/L |  | 56% (181) | 1 | REFERENCE |  |
|  | ≥10 mmol/L |  | 44% (142) | 1.65 | 1.07 to 2.56 | **0.03** |
| Average CBG in 1<sup>st</sup> 72hr after COVID-19 diagnosis |  | 306 |  |  |  |  |
|  | <10 mmol/L |  | 64% (197) | 1 | REFERENCE |  |
|  | ≥10 mmol/L |  | 36% (109) | 1.93 | 1.22 to 3.11 | **0.008** |

62% of our cohort were from non-White ethnic backgrounds, including 17% Black and 11% South Asian. The commonest co-morbidities were hypertension (47%), hypercholesterolaemia (43%), diabetes (38%) and ischaemic heart disease (IHD, 16%).

By comparison, amongst the patients treated for COVID-19 who had diabetes, the average age was 68.5±14.6 years and 66% were men. Average weight was 83.5 (±18.9) kg for men and 77.8 (±24.5) kg for women. In this cohort of patients with diabetes, 72% were non-White, including 20% Black and 14% South Asian; and a past medical history of hypertension was present in 70%, IHD in 28% and hyperlipidaemia in 51%. The use of glucose lowering medications in this population at the time of diagnosis of COVID-19 is summarised in Table 1. Average HbA1C was 64.7 (±21.7) mmol/mol (8%), 27% were taking an angiotensin converting enzyme (ACE) inhibitor and 63% were taking a statin.

### FACTORS AT THE TIME OF DIAGNOSIS OF COVID-19

Table 2 summarises the differences in biochemical measurements and clinical measurements between patients who fulfilled the primary outcome compared with patients who survived and were not admitted to ICU; and between patients with a diagnosis of diabetes and those without diabetes. As expected, biochemical parameters established to be associated with severe infection were significantly higher in the group who fulfilled the primary outcome of death/ICU admission, including C-reactive protein (CRP), d-dimers, procalcitonin, brain natriuretic peptide (BNP) and ferritin. For both patients with diabetes and those without, a capillary blood glucose (CBG) below 10 mmol/L at presentation was associated with a lower risk of poor outcome. In the cohort without diabetes, this association was sustained after correction for age and gender as described elsewhere^14^, however this was not sustained on multivariate analysis in our cohort (see below).

**Table 2.**
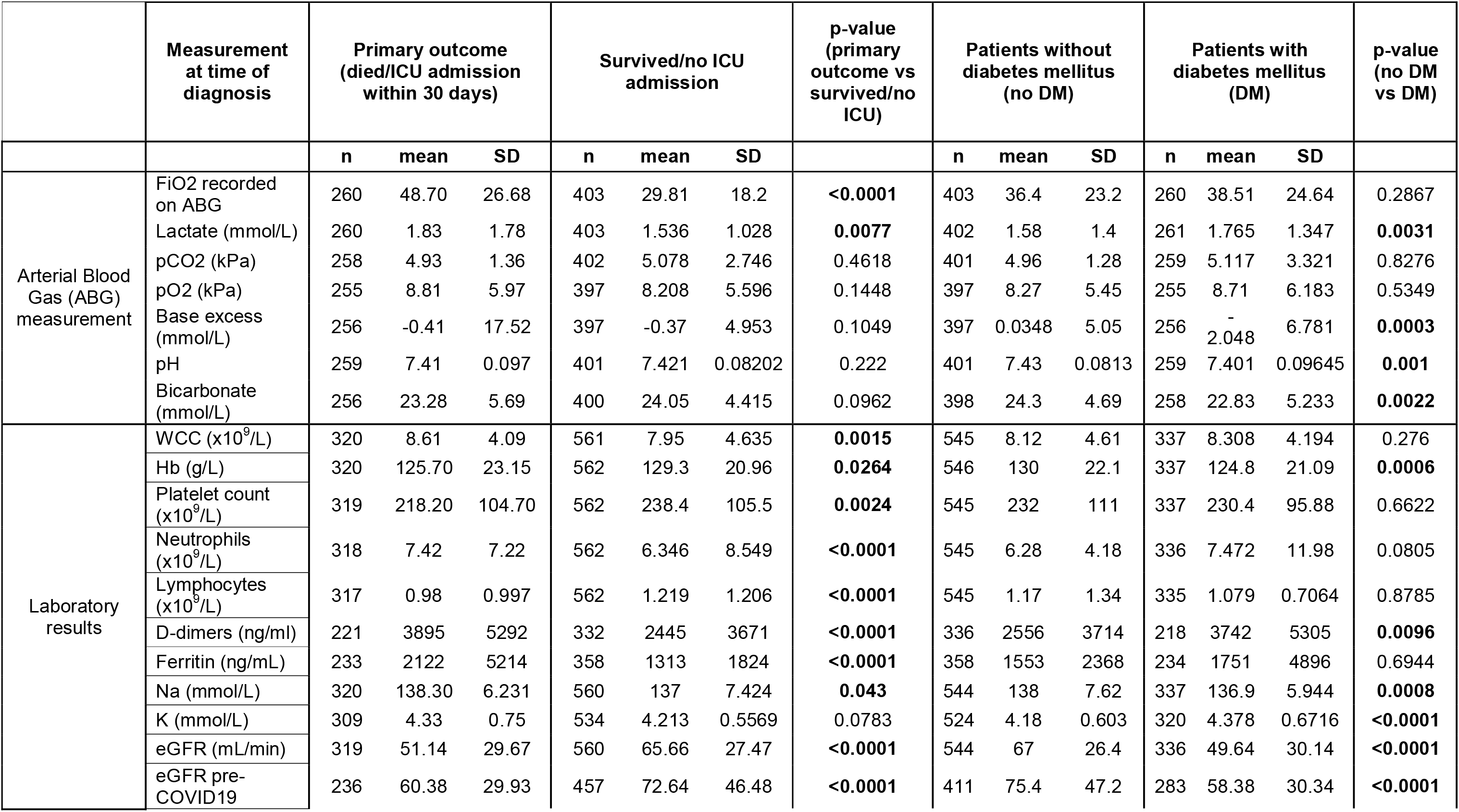
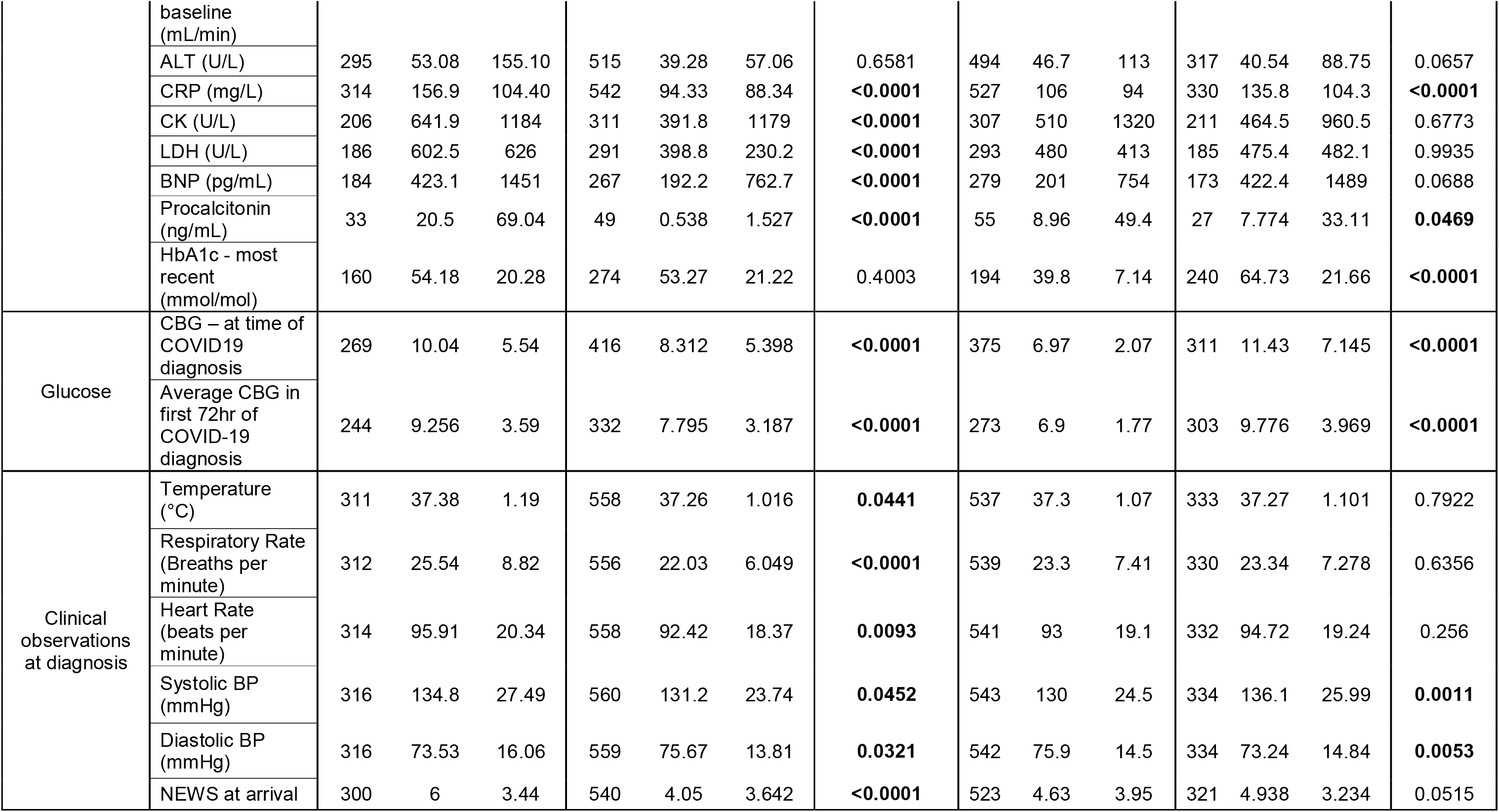
- Biochemical Measurements and Clinical Observations for 889 patients admitted with COVID-19. Unadjusted comparisons are shown for patients who met the primary outcome (death or ICU admission within 30 days of diagnosis) against o survived and did not have an ICU admission (columns 3-5); and for patients with diabetes against those without (columns 6-8). n = number of for whom that data set is available. Note the total number of patients arriving at the primary outcome was 323 and the total number of patients with diabetes mellitus (DM) was 327. ABG = arterial blood gas, ALT = alanine aminotransferase, BNP = brain natriuretic peptide, BP = blood pressure, apillary blood glucose, CRP = C-reactive protein, CK = creatinine kinase, eGFR = estimated glomerular filtration rate, FiO2 = fraction of inspired Hb = haemoglobin, HbA1c = glycated haemoglobin, K = potassium, LDH = lactate dehydrogenase, Na = sodium, NEWS = National Early Warnin CC = white cell count.

Amongst patients with diabetes, those with more severe infection had significantly lower arterial blood gas pH values, lower bicarbonate levels and higher serum potassium levels (although still, on average, within the laboratory reference range) at the time of COVID-19 diagnosis. This hints at the possibility that patients in the primary outcome group were more likely to have diabetic metabolic emergencies at presentation, similar to what has been reported by other groups^15,16^. Indeed, in our cohort there were 59 episodes of diabetic ketoacidosis (DKA) associated with COVID-19 during the data collection period.

Chest radiographs on diagnosis were reported by in-house radiologists on a five-point severity scale (1 = normal, 5 = widespread, dense bilateral infiltrates). 60% of patients who survived without ICU admission had classic patchy ground glass changes, with a median severity score of 2 out of 5. Amongst those who died/were admitted to ICU, 72% had classic radiological features with a median severity score of 3. These proportions were similar amongst the patients with diabetes.

### CLINICAL FEATURES IN THE WHOLE COHORT ASSOCIATED WITH THE PRIMARY OUTCOME (DEATH OR ICU ADMISSION WITHIN 30 DAYS OF COVID-19 DIAGNOSIS)

On univariate analysis, the risk of the primary outcome (death or ICU admission within 30 days of COVID-19 diagnosis) was significantly higher in men and older age groups (Table 1). Several co-morbidities (i.e. hypertension, ischaemic heart disease and heart failure) were also associated with an increased risk of poor outcome. Patients taking an ACE inhibitor had an odds ratio of 1.57 (CI 1.10 to 2.22, p=0.013) for the primary outcome, but an increased risk was not statistically evident with patients on angiotensin receptor blocker (ARB) drugs. The Clinical Frailty Scale (CFS) is widely used in the UK and scores range from 1 (very fit) to 9 (terminally ill)^17^. Here we show that the CFS was a good predictor of poorer outcomes, with patients scoring ≥7 having an odds ratio of 2.34 (1.46 to 3.76, p=0.0004) for death/ICU admission compared with those scoring 1-2. Similarly, an estimated glomerular filtration rate (eGFR) at the time of COVID-19 diagnosis of <45 mL/min was associated with a 3 to 4-fold increased risk of death/ICU admission. Weight >80kg did not reach significance for predicting poorer outcomes.

After removing datasets where >5% datapoints were missing (i.e. 170 patients), we ran an unselected multivariate logistic regression analysis of 37 regressors (variables) against the primary outcome on 719 patients (Table 3). Age and gender retained significance as independent variables. As well as reporting p-values for the significance of the independent contribution of a given variable, we have also calculated marginal effect sizes. This percentage value is a measure of the altered risk of primary outcome (death/ICU admission) as the given variable changes. The marginal effect of male gender was +9.3%, i.e. all other factors being the same, the risk of primary outcome was 9.3% higher for men than women. The CFS was also found to be a significant independent predictor of death/ICU admission. Each unit rise in score (from 1-9) was associated with a 3.1% increased risk of death/ICU admission, all other things being equal.

**Table 3.** - Unbiased Multivariate Logistic Regression Analysis of 37 Regressors (variables) against the Primary Outcome of Death/ICU Admission within 30 Days of COVID-19 Diagnosis (n=719 patients) This is an unselected multivariate logistic (Logit) analysis of all variables that were collected for patients admitted with swab positive COVID-19. 719 patients are included with 37 variables – with the only exclusions being those patients/variables for which ≥5% data points were unknown. For categorical variables, a positive “estimate” indicates an increased risk of the primary outcome (death or ICU admission) with that variable present, and a negative estimate indicates a reduced risk of the primary outcome if that variable is present. The p-value is a measure of the confidence of that given variable being an independent predictor of the primary outcome corrected for all of the other regressors listed. For continuous variables, a positive “estimate” indicates an increasing risk of the primary outcome as the variable increases. Since in logistic regressions estimated coefficients cannot be interpreted as a measure of the contribution of the effect, we have also calculated marginal effects. A positive marginal effect indicates that an increase in that variable (as defined in Table 2) is associated with a fully adjusted increased risk of the primary outcome. The converse applies for negative marginal effects. For categorical variables, the marginal effect indicates the percentage increased risk of the primary outcome, if that variable exists. So, for example, all other variables being equal, men have a 9.3% increased risk of death or ICU admission than women. eGFR = estimated glomerular filtration rate. #The effect of serum sodium was skewed by patients with serum sodium >145 mmol/L.

| Regressor | Estimate | SE | p-value | Marginal Effect (%) |
| --- | --- | --- | --- | --- |
| Age | 0.029 | 0.011 | **0.005** | **0.5** |
| Male gender | 0.553 | 0.256 | **0.031** | **9.3** |
| Ethnicity - South Asian | -0.321 | 0.441 | 0.466 | -5.4 |
| Ethnicity - Black | -0.296 | 0.360 | 0.411 | -5.0 |
| Ethnicity - White | -0.179 | 0.272 | 0.510 | -3.0 |
| Diabetes mellitus | 0.099 | 0.265 | 0.709 | 1.7 |
| Stroke | -0.457 | 0.348 | 0.189 | -7.7 |
| Hyperlipidaemia | 0.117 | 0.277 | 0.673 | 2.0 |
| Ischaemic heart disease | 0.596 | 0.364 | 0.102 | 10.1 |
| Heart Failure | 0.046 | 0.408 | 0.910 | 0.8 |
| Hypertension | 0.207 | 0.285 | 0.468 | 3.5 |
| Chronic obstructive pulmonary disease | -0.051 | 0.384 | 0.893 | -0.9 |
| Active cancer | -0.135 | 0.428 | 0.752 | -2.3 |
| Statin | -0.310 | 0.288 | 0.282 | -5.2 |
| Angiotensin converting enzyme inhibitor | 0.351 | 0.329 | 0.285 | 5.9 |
| Angiotensin II receptor blocker | 0.145 | 0.366 | 0.691 | 2.5 |
| Antiplatelet drug | -0.616 | 0.319 | **0.053** | **-10.4** |
| Clinical Frailty Scale | 0.183 | 0.078 | **0.019** | **3.1** |
| White cell count | 0.031 | 0.085 | 0.720 | 0.5 |
| Haemoglobin | -0.008 | 0.006 | 0.182 | -0.1 |
| Platelet count | -0.004 | 0.001 | **0.001** | **-0.1** |
| Neutrophils | 0.009 | 0.086 | 0.920 | 0.1 |
| Lymphocytes | -0.011 | 0.229 | 0.963 | -0.2 |
| Serum sodium | 0.072 | 0.023 | **0.002<sup>#</sup>** | **1.2** |
| Serum potassium | 0.202 | 0.194 | 0.300 | 3.4 |
| eGFR at diagnosis | -0.006 | 0.005 | 0.287 | -0.1 |
| C-reactive protein | 0.002 | 0.001 | 0.132 | 0.0 |
| Temperature | -0.012 | 0.113 | 0.913 | -0.2 |
| Respiratory rate | 0.034 | 0.021 | 0.103 | 0.6 |
| Heart rate | 0.006 | 0.008 | 0.453 | 0.1 |
| Systolic blood pressure | 0.010 | 0.006 | 0.088 | 0.2 |
| Diastolic blood pressure | -0.002 | 0.010 | 0.831 | 0.0 |
| National Early Warning Score | -0.015 | 0.055 | 0.790 | -0.2 |
| Inspired oxygen delivered at diagnosis | -0.012 | 0.006 | **0.046** | **-0.2** |
| Oxygen saturations at diagnosis | -0.032 | 0.018 | 0.079 | -0.5 |
| Maximum inspired oxygen required during admission | 0.068 | 0.007 | **<0.0001** | **1.2** |

The association between ethnicity and poor outcome was not sustained on multivariate analysis in our cohort. None of the individual co-morbidities we collected data on (i.e. diabetes, hypertension, hyperlipidaemia, IHD, cerebrovascular disease, heart failure, chronic obstructive airway disease (COPD) and cancer) were found to be independent predictors of mortality/ICU admission in this analysis, including diabetes.

Taking an antiplatelet drug was significantly and independently associated with a 10% lower risk of death and/or ICU admission (Table 3). Related to this, platelet count was a significant predictor of poor outcome. Our data suggest that every reduction in platelet count of 100 x10^9^/L is associated with a 10% risk of death/ICU admission. This association remained after removal of patients with platelet counts below 50 x 10^9^/L (the majority of whom had active, often haematological, malignancies). Higher serum sodium at the time of COVID-19 diagnosis was also found to be a strong independent predictor of poor outcome, however this association disappeared after removing patients with high sodium (i.e. >145 mmol/L). As a sensitivity analysis, we performed a regularised regression (Smoothly Clipped Absolute Deviation - SCAD)^18-20^, which selects the truly significant variables in the multivariate regression without the limitations of a stepwise approach (see Supplementary Methods). This SCAD analysis identified 23 variables driving the variance. It supported the findings of the unrestricted multivariate regression (Supplementary Table 1).

### CLINICAL FEATURES IN PATIENTS WITH DIABETES MELLITUS ASSOCIATED WITH THE PRIMARY OUTCOME

Unrestricted multivariate logistic analysis with >95% complete datasets was possible on 278 patients with diabetes (Table 4). This excluded HbA1C from the analysis due to inadequate data. However, multivariate analysis on 197 patients for HbA1C did not produce a significant interaction with the primary outcome (p>0.9) and neither was it a significant factor on univariate analysis (Table 1). On the unrestricted multivariate analysis of patients with diabetes, no medication type was significantly associated with poorer outcomes. Of all the associated co-morbidities examined, ischaemic heart disease had a highly significant 33% marginal effect for increased likelihood of death/ICU admission. Other factors significantly and independently associated with poor outcome in patients with diabetes were age, male gender and lower platelet count (Table 4). This was corroborated on SCAD analysis (Supplementary Table 2)

**Table 4.**
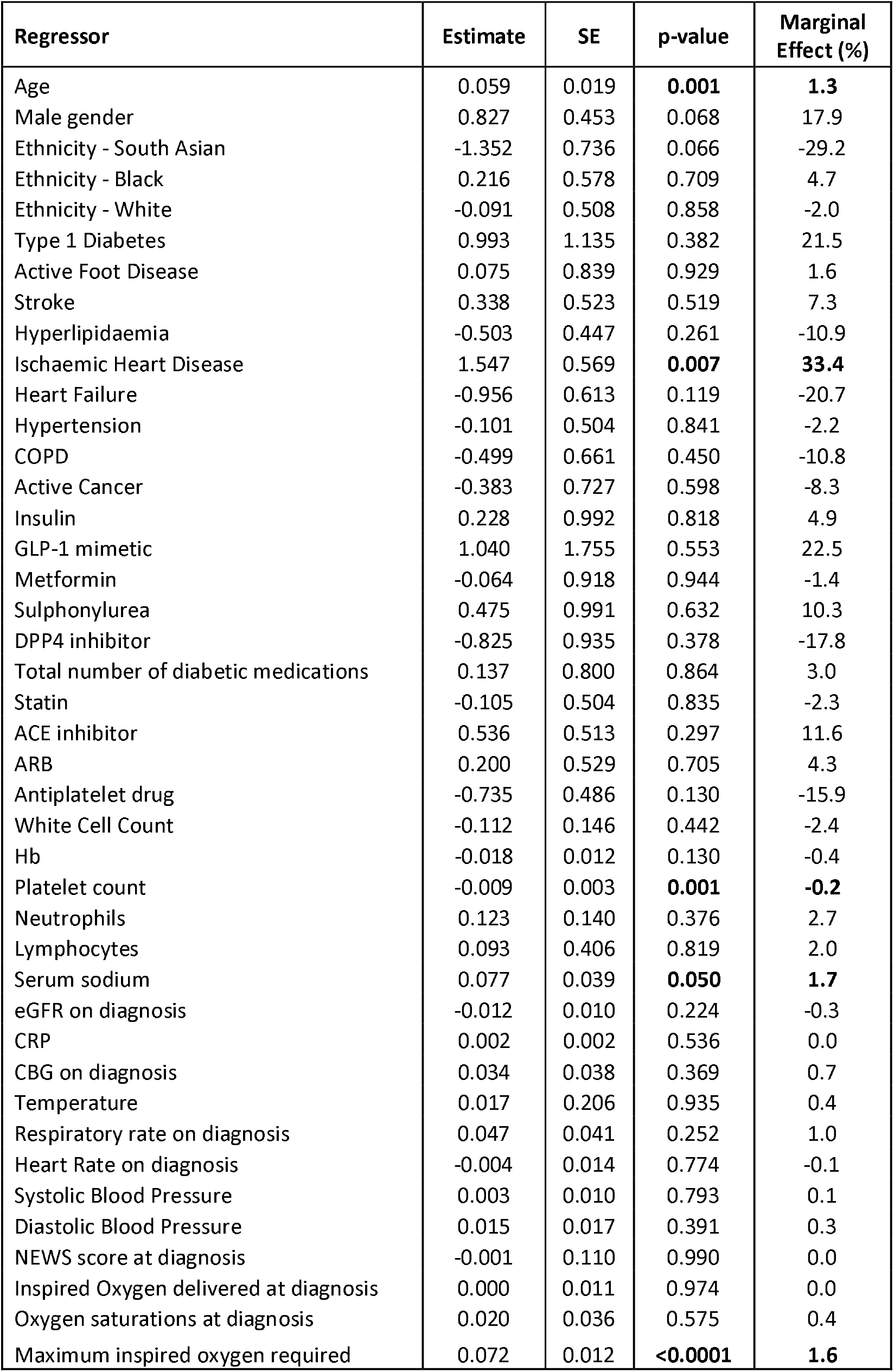
- Unbiased Multivariate Logistic Regression Analysis of 42 Regressors (variables) against the Primary Outcome of Death/ICU Admission within 30 Days of COVID-19 Diagnosis in patients with Diabetes Mellitus (n=268 patients) This is an unselected multivariate logistic (Logit) analysis of all variables that were collected for patients admitted with swab positive COVID-19 who had diabetes mellitus, as applied to the primary outcome of death or ICU admission within 30 days. 268 patients are included with 42 variables – with the only exclusions being those patients/variables for which ≥5% data points were unknown. For this reason, HbA1C is not included as a regressor as it would have reduced the number of patients included to 168 – although of note in that regression, HbA1c did not survive multiple correction (and neither did body weight). For categorical variables, a positive “estimate” indicates an increased risk of the primary outcome (death or ICU admission) with that variable present, and a negative estimate indicates a reduced risk of the primary outcome if that variable is present. The p-value is a measure of the confidence of that variable being an independent predictor of the primary outcome corrected for all of the other regressors listed. For continuous variables, a positive “estimate” indicates an increasing risk of the primary outcome as the variable increases. Since in logistic regressions estimated coefficients cannot be interpreted as a measure of the contribution of the effect, we have also calculated marginal effects along with their standard errors. A positive marginal effect indicates that an increase in that variable (as defined in Table 2) is associated with a fully adjusted increased risk of the primary outcome. The converse applies for negative marginal effects. For categorical variables, the marginal effect indicates that percentage increased risk of the primary outcome, if that variable exists. So, for example, patients with diabetes (all other things being equal) have a 33% increased risk of death/ICU if they have ischaemic heart disease. eGFR = estimated glomerular filtration rate.

### STRATIFIED MARGINAL EFFECT SIZE FOR THE SIGNIFICANT PREDICTORS THAT SURVIVED MULTIPLE LOGISTIC REGRESSION: PLATELET COUNT AND THE CLINICAL FRAILTY SCALE (CFS)

It has been suggested that the CFS is a reasonable measure to include in decision-making tools for escalation to ICU care^21^. However, it was developed for use in people aged >65 years^17^, and has not been validated in the general population as a tool for guiding escalation decisions. Therefore, we stratified the marginal effect of the CFS score as an independent predictor of poor outcome in COVID-19 by age group (Figure 2a). This confirms that the CFS is a useful predictor of primary outcome in older age groups. Similarly, the independent marginal effect of platelet count tends to increase with age (Figure 2b) but there was no interaction with the use of antiplatelet drugs (p>0.9).

**Figure 2:**
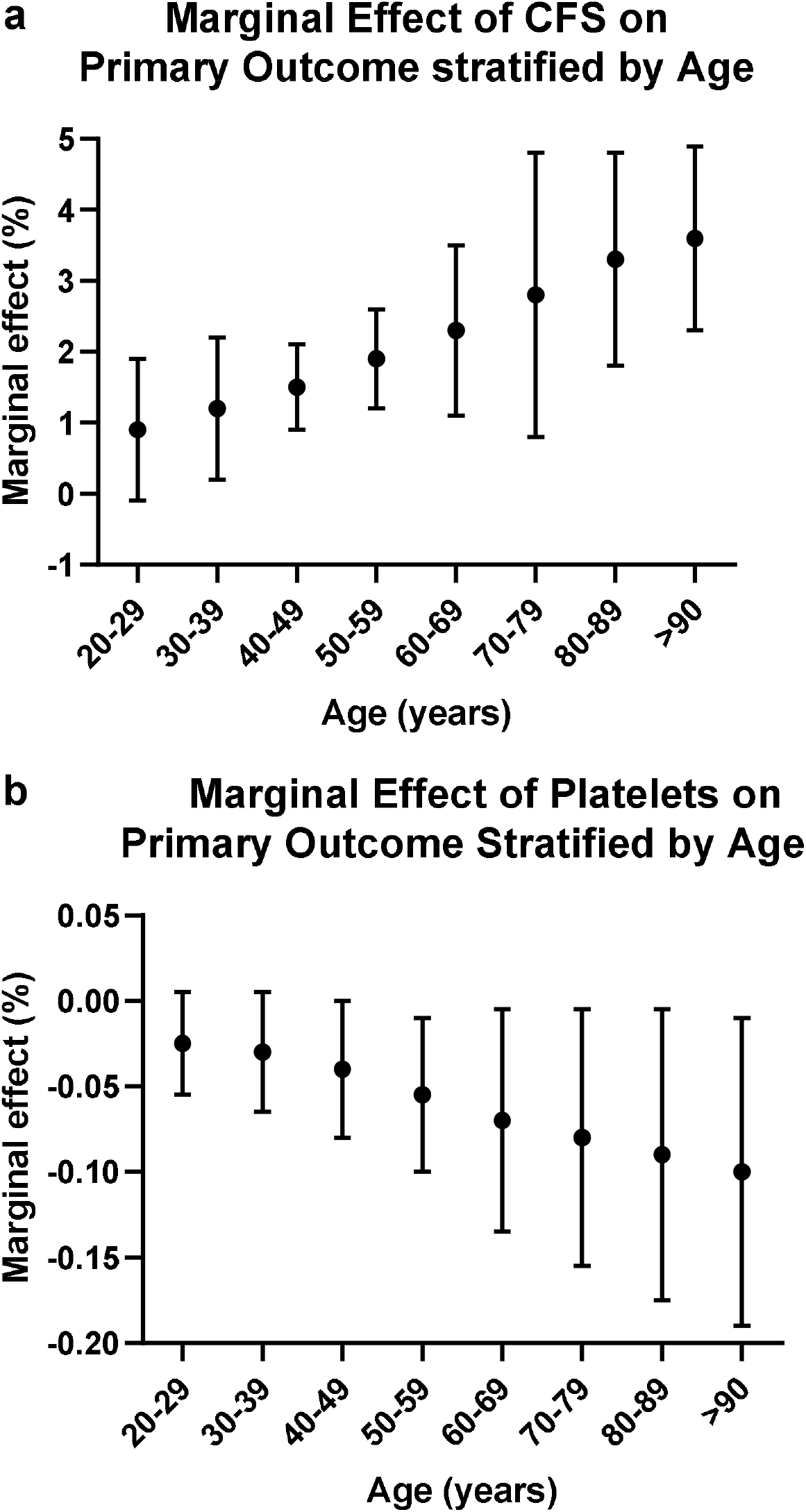
On unbiased univariate analysis of 798 patients admitted with COVID-19, the Clinical Frailty Scale (CFS) was a significant independent predictor of poor outcomes, with a marginal effect size of 3% for every increase in score between 1-9 (see Table 3). We stratified the marginal effect by age bands. There was a trend (Figure 2a) for the marginal effect of the CFS on the primary outcome to be greater in the older population. Similarly, there was a trend for the independent marginal effect of platelet count on the primary outcome to increase with age (Figure 2b), but there was no interaction with the marginal effect size of platelet count on outcomes as stratified by the use of antiplatelet drugs (data not shown).

## Discussion

Here we report on a large cohort of patients diagnosed with COVID-19 over a 6-week period in a multi-site NHS trust in London. In our cohort, multivariate analysis revealed male gender, increased age, increased frailty and lower platelet count were independently associated with increased risk of ICU admission and/or death within 30 days of COVID-19 diagnosis, while taking antiplatelet medication was associated with a lower risk of poor outcome. Furthermore, within the subset of our cohort with diabetes (96% Type 2 and 4% Type 1), multivariate analysis demonstrated male gender, pre-existing ischaemic heart disease, advancing age and lower platelet count were associated with increased risk of ICU admission and/or death.

In general, patients with diabetes are at increased risk of hospitalisation. Prior to the COVID-19 pandemic, data from the National Diabetes Inpatient Audit indicate the prevalence of diabetes amongst hospital inpatients in England and Wales is 18%^22^. Using data from a UK primary care database, Barron et al. reported that people with both Type 1 and Type 2 diabetes had multiply-adjusted increased odds of dying in hospital with COVID-19 compared to those without diabetes^23^. Our results show that people with diabetes are at increased risk of severe or life threatening COVID-19, although this was driven by its tendency to co-exist with other conditions, particularly ischaemic heart disease. The relative proportions of patients admitted with COVID-19 with Type 1 and Type 2 diabetes were similar to the population we serve, suggesting no difference in susceptibility based on diabetes type.

Hyperglycaemia is a modifiable factor that may influence outcome in COVID-19, especially in people with diabetes. In our cohort, recent HbA1c was not a significant predictor of poor outcome, which is similar to some other cohort studies^13^. In patients with diabetes (and in patients without diabetes), our data demonstrate that blood glucose ≥10 mmol/L (at the time of COVID-19 diagnosis and on average during the 72 hours following COVID-19 diagnosis) is associated with increased risk of death/ICU admission. Similar findings have been reported by groups from France^13^ and China^12,14^. This association is not sustained on multivariate analysis in our patients with diabetes and there is no prospective study to address whether maintaining blood glucose <10 mmol/L would improve outcomes for patients with diabetes with COVID-19. Furthermore, maintaining blood glucose <10 mmol/L may become even more challenging to achieve as the use of dexamethasone in the management of severe COVID-19 becomes more widespread, following publication of beneficial reports of its use in this context^24^.

It is unsurprising that diabetes should be associated with a poorer outcome as it is in many infections, often attributed to the relative immunocompromise associated with hypergycaemia^25^. It is now well established that patients with more severe manifestations of COVID-19 (including those that need to be escalated to ICU care and those that die) are much more likely to have a diagnosis of diabetes than those who are documented as having a mild form of the infection^4,23,26^. Some people have recommended that patients with diabetes need to be more actively shielded, and diabetes may be associated with a higher risk of viral infection^27^. However, it is important to note that here we show that diabetes, in its own right, is not a major factor contributing to the risk of death/ICU admission, but rather its association with other consequences of the metabolic syndrome confers a higher risk of a poor outcome. For instance, all other things being equal, patients (in our cohort) with diabetes have a 33% increased risk of death/ICU admission if they also have ischaemic heart disease.

In line with our observations that it is multimorbidity *per se* and not any particular diagnosis that confers a strong increased risk of poorer outcomes in COVID-19 infection, we show that the CFS is a robust, independent predictor of poor outcome on multivariate analysis. This is consistent with (COVID-19 and non-COVID-19) studies that show that as the CFS increases, the likelihood of mortality increases^21,28^. However, we have also shown that the CFS is much less useful in younger age groups. We chose death or ICU admission as our primary outcome measure, as this incorporated all patients with severe (life threatening) COVID-19. Our cohort also included those who, with unfavourable chances of tolerating and surviving invasive ventilation, would not have been admitted to ICU due to pre-existing multimorbidity.

We report a strongly significant and independent risk of death/ICU admission as platelet count decreases. The inverse relationship between platelet count and risk of death/ICU admission with COVID-19 has also been reported by several other studies^26,29,30^. As microvascular and macrovascular thrombosis is increasingly being reported as a feature of severe COVID-19^31-33^, reduced platelet count may reflect consumptive coagulopathy (d-dimers were significantly higher in the primary outcome group), possibly in conjunction with direct effects on thrombopoiesis or platelet survival. There is no international consensus yet on the use of anticoagulation in COVID-19. We show that patients on antiplatelet drugs were more likely to survive (all other things being equal), even though ischaemic heart disease (the most common indication for their use) was an independent predictor of poorer outcome.

Strengths of our study include a diverse population of patients, and in-depth characterisation, including CFS and numerous pre-hospital and presentation factors. We selected a statistical approach focussed on a primary outcome measure with no selection bias for the multivariate analysis that produced intuitive results, which survived robust sensitivity analysis.

Some of our findings are at odds with other reports. For example, we report no particular association of Black or South Asian ethnicity as an independent risk factor for poorer outcome. However, in 34% of our cohort, ethnicity was reported as mixed, other or unknown, which may have accounted for the disparity between our results and reports that suggest people with non-White ethnicity have poorer outcomes^34^. Furthermore, in our clinical experience we noted a strong interaction between ethnicity and socioeconomic strata, hence job type – with many lower paid jobs associated with the inability to work from home. In the UK, people of Black and South Asian ethnicities are also more likely to work in frontline healthcare roles^35^. Socioeconomic data, including job family, should be included in future analyses. It is worthy of note that patients of Black ethnicity were over-represented in our cohort compared to population levels in the community we serve (Black: 17% in our cohort, 7-12% in the community; and South Asian: 11% in our cohort, 9-12% in the community)^36^. It remains to be understood to what extent this is a reflection of increased risk of infection due to socioeconomic factors as opposed to other factors.

We were unable to include BMI as a regressor in this dataset as height is not routinely recorded in our Trust. When body weight was included in our multivariate analysis, it did not contribute independently and was removed due to insufficient patients with accurate data. Other studies have concluded that BMI is an important predictor of poorer outcomes, including for patients with diabetes. This concurs with our message that it is not a particular disease *per se* but rather a constellation of factors (typified by the metabolic syndrome) that confers risk. Estimated glomerular filtration rate (eGFR) was also not an independent predictor for poor outcome in patients with diabetes.

Our data suggest that diabetes should not be considered differently to other common co-morbidities in the risk assessment for patients with COVID-19. The tendency for diabetes to cluster with more significant predictors of poor outcome such as ischaemic heart disease is important to recognise, in addition to other independently associated risk factors such as age, male gender and lower platelet count. This study contributes further to understanding the drivers of poor outcomes in patients with diabetes admitted to hospital with COVID-19.

## Conclusions

This study contributes to the pool of knowledge of the determinants of poor outcomes with COVID-19. Male gender, ischaemic heart disease, advancing age, multi-morbidity (as crystallised in the Clinical Frailty Scale) and lower platelet count are important predictors of poor outcome in patients with severe COVID-19 (both in patients with diabetes and those without diabetes). Antiplatelet medications confer an advantage. There is no clear evidence that dysglycaemia drives the increased risk of death/ICU admission amongst patients with diabetes, but rather the association of diabetes with other common medical conditions confers excess risk of poor outcomes.

## Methods

### STUDY SETTING

We performed this retrospective cohort study at Imperial College Healthcare NHS Trust (ICHNT), which includes three hospitals admitting patients with COVID-19 (Charing Cross Hospital, Hammersmith Hospital and St Mary’s Hospital). All patients who had a nasopharyngeal swab taken to determine SARS-CoV-2 infection between 9^th^ March 2020 and 22^nd^ April 2020 were identified on an automated search of the laboratories’ computer systems. COVID-19 diagnosis was determined by the presence of SARS-CoV-2 infection as evidenced by a positive reverse transcription-polymerase chain reaction (RT-PCR) result from a nasopharyngeal swab. RT-PCR was performed on nasopharyngeal swabs by NorthWest London Pathology staff in the laboratories in the constituent hospitals of ICHNT.

Excluded from data collection were: all patients with negative SARS-CoV-2 swab results, patients who did not have an Emergency Department attendance and/or an inpatient admission within ICHNT, and all patients aged less than 18 years (Figure 1). This study was approved by the ICHNT Clinical Audit Office, was performed in accordance with the Declaration of Helsinki and UK Data Protection legislation. For patients with more than one positive swab result or multiple hospital admissions during the data collection period, the date of the earliest swab result and the most significant hospital admission in terms of COVID-19 severity, respectively, was used for data collection.

### DATA COLLECTION

Data collection was performed by physicians working in ICHNT. Hospital numbers of patients with positive swab results were checked against electronic health records held on the ICHNT computer system (Cerner Corporation, Kansas City, USA). Data is uploaded onto Cerner manually by health practitioners and administrators to record and store patient notes, prescribe and dispense medication; as well as store investigation results and clinic letters. Furthermore, Cerner is connected to the National Health Service (NHS) Spine, which synchronises other data (such as date of death).

Demographic, clinical and biochemical data (on the date of admission/Emergency Department attendance or on the date of the first positive SARS-CoV-2 swab result if the patient had been admitted for ≥1 week), as well as outcome data were collected for all eligible patients up to and including 20^th^ June 2020 to ensure there was a minimum of 30 days of follow-up after the date of COVID-19 diagnosis to eliminate the issue of censoring.

Data collected included: demographic and clinical characteristics (age, gender, ethnicity, latest body weight), past medical history, specific comorbidities and medications on admission. Clinical details about diabetes were noted including: classification of diabetes, duration of diabetes and hypoglycaemic medications. A score using the Clinical Frailty Scale (CFS)^17^ was assigned to every patient. Haematological and biochemical data for the day of COVID-19 diagnosis included full blood count, routine biochemistry and arterial blood gas results. Average capillary blood glucose values were recorded for the first three days following COVID-19 diagnosis and the most recent HbA1c result (within 6 months) was also recorded.

### STATISTICAL ANALYSIS

The composite primary outcome was admission to ICU or death within 30 days of COVID-19 diagnosis, and data on the primary outcome was collected for all patients. This was chosen to capture patients with severe and/or life-threatening COVID-19. Univariate and then unrestricted multivariate logistic regression analyses were performed to look for factors associated with an increased risk of the primary outcome (i.e. death or ICU admission within 30 days of COVID-19 diagnosis). Unlike a Cox proportional hazards approach, this analysis focusses more on “if” a patient gets severely ill rather than survival duration.

The first stage of analysis was descriptive. Quantitative data are expressed as mean ± SD. Categorical variables were given as percentage (number) of participants. Univariate logistic regression models were used to calculate odds ratios (OR) associated with the primary outcome. Comparison between two groups was analysed using Student’s t-tests (normally distributed data) or Mann-Whitney U test (non-normally distributed data) for continuous variables. Comparison of categorical variables was analysed by Fisher’s exact test or χ^2^ test. A difference with a two-sided α < 0.05 was considered statistically significant.

Multivariable logistic regression was applied to assess the independent association of the primary outcome with all the clinical and biological features included in the study. For the top level (unrestricted) analysis only datasets with <5% missing values were included, leaving a total n number of 716 patients and 37 predictors (variables). No data points were imputed. Data was winsorized (at 1% level) to neutralise the effects of the most extreme values. Following this first step, a regularised regression (Smoothly Clipped Absolute Deviation - SCAD) analysis^18-20^ was performed to independently account for the over-dimensionality of the unrestricted multivariate. In brief, this sensitivity analysis allows unbiased selection of the most relevant and important predictors and controlling the errors involved in variable selection, without the need to re-estimate the model in a stepwise fashion. Results are presented as the estimate parameter, the p-value of the association and the marginal effect size.

Next, only patients with diabetes were included in multivariate and regularised regression analyses, to look for elements of their clinical presentation that were associated with poorer outcome with COVID-19. Finally, marginal effects for the most significant (independent) risk factors were sub-stratified by age (for the CFS) and the presence or absence of ischaemic heart disease or usage of antiplatelet agents (for platelet counts). All statistical analyses (VS and WD) were performed on GraphPad Prism V8.1 (descriptive statistics) and custom written MaTLAB scripts (multivariate analyses). A detailed description of statistical methods can be found in the Supplementary Methods.

## Data Availability

Data requests should be made to the corresponding author.

## Acknowledgments

The Section of Endocrinology and Investigative Medicine is funded by grants from the MRC, BBSRC, NIHR and is supported by the NIHR Biomedical Research Centre Funding Scheme. The views expressed are those of the authors and not necessarily those of the MRC, BBSRC, the NHS, the NIHR or the Department of Health and Social Care. CI is funded by an Imperial-BRC IPPRF Fellowship, JK is funded by an NIHR Doctoral Research Fellowship (DRF-2017-10-042), SM is funded by a Future Leaders Mentorship Award from the European Association for the Study of Diabetes, TT is funded by the MRC and NIHR, VS is funded by a Harry Keen Diabetes UK Fellowship.

## Author Contributions

Conceptualization, CI, TT, NH and VS.; Methodology, CI, AA, DH, TT, NH and VS; Investigation, CI, AA, WY, OI, JK, RS, EW, CS, NA, HB, NB, SB, ML, SP, ES, ET, AQ, MA, DH, NO, VB, SM, TT, NH and VS.; Formal analysis, WD, VS; Data curation, CI, AA, SM, TT, NH and VS; Writing—original draft preparation, CI and VS.; Writing—review and editing, CI, WD, AA, WY, OI, JK, RS, EW, CS, NA, HB, NB, SB, ML, SP, ES, ET, AQ, MA, DH, NO, VB, SM, TT, NH and VS.; Visualization, CI, WD and VS. All authors have read and agreed to the published version of the manuscript.

